# Comparison of Surgical versus Transcatheter Pulmonary Valve Replacement Outcomes in Real-World Clinical Practice

**DOI:** 10.1101/2025.08.01.25332785

**Authors:** Azka Naeem, Olayiwola Bolaji, Muhammad Hashim Khan, Krishna Vamsy Polepalli, Abdullahi Oshioke Oseni, Jaffar M. Khan, Omar Khalique

## Abstract

**Background:** Pulmonary valve dysfunction commonly necessitates intervention in congenital heart disease patients. While surgical pulmonary valve replacement (SPVR) remains the traditional approach, transcatheter pulmonary valve replacement (TPVR) has emerged as a less invasive alternative. This study aims to compare real-world outcomes between these approaches using large-scale, multi-institutional data.

**Methods:** This retrospective cohort study utilized the TriNetX Research Network to identify patients undergoing SPVR or TPVR between January 2010 and December 2024. After propensity score matching, 5,322 patients were included in each cohort. Primary outcomes included pulmonary hypertension, all-cause mortality, and major adverse cardiovascular events (MACE). Secondary outcomes comprised cardiogenic shock, permanent pacemaker implantation, mechanical ventilation, acute kidney injury, and endocarditis. Outcomes were assessed at 3 months, 6 months, 1 year, 3 years, and 5 years post-procedure.

**Results:** At 5-year follow-up, no significant differences were observed between SPVR and TPVR groups in rates of pulmonary hypertension (HR 0.912, 95% CI 0.773-1.077, p=0.278), all-cause mortality (HR 0.963, 95% CI 0.851-1.089, p=0.545), or pacemaker requirement (HR 0.949, 95% CI 0.850-1.059, p=0.346). However, SPVR was associated with higher risk of mechanical ventilation (HR 1.140; 95% CI: 0.963-1.349, p =1.29), while TPVR showed increased rates of acute kidney injury (HR 0.828, 95% CI 0.704-0.973, p=0.022) and endocarditis (HR 0.787, 95% CI 0.661-0.937, p=0.007). At shorter follow-up intervals, TPVR demonstrated significantly lower rates of cardiogenic shock (3-month: 3.5% vs. 4.5%, p=0.002), MACE (3-month: 16.5% vs. 21.9%, p<0.001), and mechanical ventilation (3-month: 23.4% vs. 30.7%, p<0.001). Multivariate Cox regression identified SPVR as an independent predictor of cardiogenic shock (HR 1.121, 95% CI 1.018-1.235, p=0.020).

**Conclusion:** This large-scale, real-world analysis demonstrates that TPVR offers significant short-term advantages with reduced procedural complications compared to SPVR, while outcomes at 5 years are largely comparable between approaches. These findings support TPVR as a viable first-line strategy in anatomically suitable candidates, particularly those at higher surgical risk, while suggesting that patient selection should carefully balance short-term procedural risk against potential long-term considerations.

## Introduction

Pulmonary valve dysfunction is a common sequela in patients with congenital heart disease, particularly following surgical repair of conditions such as Tetralogy of Fallot or pulmonary atresia. Over time, degeneration of right ventricular outflow tract (RVOT) conduits or bioprosthetic valves often necessitates re-intervention to alleviate symptoms and preserve right ventricular function [1]. Although surgical pulmonary valve replacement (SPVR) remains the traditional standard, it is associated with significant procedural morbidity and the cumulative risks of multiple sternotomies, especially in younger patients [2].

Transcatheter pulmonary valve replacement (TPVR) has emerged over the past two decades as a less invasive alternative, showing favorable outcomes in select patient populations. It offers reduced hospital stays, lower complication rates, and shorter recovery times compared to SPVR [3]. The Melody® and Sapien® valve systems have demonstrated procedural success and early hemodynamic improvement in both clinical trials and observational studies [4,5]. However, despite increasing adoption, much of the available data on TPVR outcomes stems from single-center experiences or limited registries, often lacking long-term or real-world insights. A growing concern is the incidence of post-procedural complications that may significantly impact long-term outcomes. These include the development or persistence of pulmonary hypertension, cardiogenic shock, conduction abnormalities necessitating permanent pacemaker implantation, respiratory compromise requiring mechanical ventilation, and acute kidney injury (AKI) — all of which are associated with increased morbidity and mortality [6–9]. Furthermore, comprehensive data on all-cause mortality following TPVR across broader, real-world populations remains limited.

To address these gaps, we conducted a large-scale, real-world analysis of patients undergoing TPVR using the TriNetX research network, a global health platform aggregating de-identified electronic medical records (EHRs) from multiple healthcare institutions. The primary objective of this study is to evaluate the incidence of pulmonary hypertension and all-cause mortality following TPVR. Secondary objectives include assessing the occurrence of major adverse events such as cardiogenic shock, permanent pacemaker implantation, mechanical ventilation, and acute kidney injury. This study aims to provide valuable insights into the real-world burden of complications following TPVR, thereby informing clinical decision-making and improving patient care.

## Methods

### Data Source

This retrospective cohort study utilized data from the TriNetX Research Network, a federated global health research network that provides access to harmonized, de-identified electronic health records (EHRs) from over 92 million patients across more than 170 healthcare organizations worldwide including academic medical centers, community hospitals, and specialty practices [10]. The TriNetX platform standardizes data using controlled terminologies including SNOMED CT, RxNorm, LOINC, and ICD-10-CM codes, enabling consistent analysis across contributing institutions [11]. All data were de-identified in compliance with the Health Insurance Portability and Accountability Act (HIPAA), providing only aggregated counts and statistical summaries. No protected health information was made available to the research team.

### Study Population and Design

In this study, comparative effectiveness study of patients who underwent either surgical pulmonary valve repair/replacement (SPVR) or transcatheter pulmonary valve replacement (TPVR) between January 2010 and December 2024 was conducted. Patients were identified using relevant ICD-10 procedure codes (specific codes for SPVR and TPVR).

The initial cohorts consisted of 6,333 patients in the SPVR group and 7,509 patients in the TPVR group. To minimize selection bias and control for confounding variables, propensity score matching was performed. The propensity score model included key demographic and clinical characteristics: age, sex, race, ethnicity, comorbid conditions (essential hypertension, diabetes mellitus, chronic obstructive pulmonary disease, hyperlipidemia, obesity, heart failure, chronic kidney disease), and left ventricular ejection fraction (LVEF) when available. After 1:1 propensity score matching, the final analysis included 5,322 patients in each group.

Patients with outcomes occurring prior to the index procedure were excluded from the respective outcome analyses. Additionally, patients whose index events occurred more than 20 years ago (373 in SPVR and 326 in TPVR group) were excluded from the follow-up time analysis.

### Outcomes and Follow-up

The primary outcomes of interest were all-cause mortality, major adverse cardiovascular events (MACCE: composite of death, myocardial infarction, cerebral infarction, cardiac arrest) and Pulmonary hypertension development within 5 years post-procedure (ICD-10 codes I27.0, I27.2x, I27.8x) Secondary outcomes included cardiogenic shock, requirement for permanent pacemaker, mechanical ventilation, acute kidney injury (KDIGO criteria),endocarditis

These outcomes were ascertained using standardized ICD-10-CM diagnosis codes. Patients were followed from the date of their procedure (index date) until the occurrence of the outcome of interest, death, loss to follow-up, or the end of the study period (December 31, 2024), whichever came first.

Outcomes were analyzed at multiple time points to assess both short-term and long-term differences between the two procedures: 3 months, 6 months, 1 year, 3 years, and 5 years after the index procedure.

### Statistical Analysis

Baseline characteristics of the study cohorts were described using means and standard deviations for continuous variables and counts and percentages for categorical variables. Standardized mean differences (SMD) were calculated to assess the quality of propensity score matching, with values <0.1 indicating good balance between the groups.

For each outcome at each time point, we conducted several analyses: Absolute risk differences, risk ratios, and odds ratios with 95% confidence intervals, were calculated, reporting z-statistics and corresponding p-values. Kaplan-Meier curves were generated to estimate the cumulative incidence of each outcome over time. Log-rank tests were performed to compare differences between the SPVR and TPVR groups. Cox proportional hazards models: Hazard ratios (HRs) were estimated with 95% confidence intervals. The proportionality assumption was tested using Schoenfeld residuals For outcomes that could occur multiple times in the same patient (e.g., mechanical ventilation, acute kidney injury), the mean number of instances were compared between groups using t-tests.

Multivariable logistic regression analyses were performed for pulmonary hypertension and cardiogenic shock, adjusting for potential confounders including age at index procedure, sex, diabetes mellitus, chronic kidney disease, chronic obstructive pulmonary disease, hyperlipidemia, obesity, heart failure, and left ventricular ejection fraction. Hazard ratios, coefficients, standard errors, z-statistics, p-values, and 95% confidence intervals were reported.

For all statistical tests, a two-sided p-value <0.05 was considered statistically significant. No adjustments were made for multiple comparisons, as this was considered an exploratory analysis.

### Ethical Considerations

This study was determined to be exempt from review by the Institutional Review Board as it utilized only de-identified patient data. The study was conducted in accordance with the Declaration of Helsinki and the International Conference on Harmonization Guidelines for Good Clinical Practice. As per TriNetX policy, data were only available in aggregate format, and no protected health information was accessible to the researchers, ensuring patient confidentiality..

### Reporting Guidelines

This study adheres to the Strengthening the Reporting of Observational Studies in Epidemiology (STROBE) guidelines for reporting observational studies [12] and the REporting of studies Conducted using Observational Routinely-collected health Data (RECORD) statement [13].

## Results

### Baseline Characteristics

After propensity score matching, 5,322 patients were included in each cohort. Baseline characteristics between the SPVR group and TPVR group were well-balanced across demographic, clinical, and laboratory variables (Table 1). No statistically significant differences were observed in most baseline characteristics including gender distribution, race, age, comorbidities such as chronic kidney disease, heart failure, and left ventricular ejection fraction.

**Table 1.** Baseline Characteristics After Propensity Score Matching.

| Characteristic | SPVR<br>(n=5,322) | TPVR (n=5,322) | P-value |
| --- | --- | --- | --- |
| **Demographics** |  |  |  |
| Age at index, years | 28.5± 22.7 | 28.6 ± 18.7 | 0.900 |
| Male gender, n (%) | 2,904 (54.6) | 2,887 (54.2) | 0.741 |
| White race, n (%) | 3,456 (64.9) | 3,443 (64.7) | 0.792 |
| **Comorbidities** |  |  |  |
| Hypertension, n (%) | 1,344 (25.3) | 1,467 (27.6) | 0.007 |
| Diabetes mellitus, n (%) | 597 (11.2) | 654 (12.3) | 0.086 |
| COPD, n (%) | 279 (5.2) | 296 (5.6) | 0.466 |
| Hyperlipidemia, n (%) | 851 (16.0) | 916 (17.2) | 0.090 |
| Obesity, n (%) | 367 (6.9) | 408 (7.7) | 0.126 |
| Heart failure, n (%) | 1,629 (30.6) | 1,686 (31.7) | 0.233 |
| Chronic kidney disease, n (%) | 467 (8.8) | 493 (9.3) | 0.379 |
| **Laboratory values** |  |  |  |
| LVEF (%), mean ± SD | 51.7 ± 16.6 | 50.5 ± 18.1 | 0.418 |

COPD = chronic obstructive pulmonary disease; LVEF = left ventricular ejection fraction; SD = standard deviation.

### Clinical Outcomes at 5-Year Follow-up

The median follow-up duration was 1,825 days for the SPVR group and 1,815.5 days for the TPVR group (Table 2). At 5-year follow-up, no significant differences were observed between the SPVR and TPVR groups in rates of pulmonary hypertension (HR 0.912, 95% CI 0.773-1.077, p=0.278), all-cause mortality (HR 0.963, 95% CI 0.851-1.089, p=0.545), cardiogenic shock (HR 0.912, 95% CI 0.681-1.221, p=0.535), requirement for pacemaker (HR 0.949, 95% CI 0.850-1.059, p=0.346), or MACE (HR 0.937, 95% CI 0.813-1.079, p=0.364).

**Table 2.** Clinical Outcomes at 5-Year Follow-up.

| Outcome | SPVR<br>(n=5,322) | TPVR<br>(n=5,322) | Risk Ratio<br>(95% CI) | Hazard<br>Ratio (95%<br>CI) | P-value |
| --- | --- | --- | --- | --- | --- |
| **Pulmonary**<br>**hypertension, n (%)** | 278/4,265<br>(6.5) | 281/4,273<br>(6.6) | 0.991 (0.844-1.163) | 0.912 (0.773-1.077) | 0.278 |
| **All-cause mortality,**<br>**n (%)** | 509/5,293<br>(9.6) | 497/5,289<br>(9.4) | 1.023 (0.910-1.151) | 0.963 (0.851-1.089) | 0.545 |
| **Cardiogenic shock, n**<br>**(%)** | 88/4,899<br>(1.8) | 92/4,962<br>(1.9) | 0.969 (0.725-1.294) | 0.912 (0.681-1.221) | 0.535 |
| **Pacemaker**<br>**requirement, n (%)** | 633/4,736<br>(13.4) | 647/4,788<br>(13.5) | 0.989 (0.893-1.095) | 0.949 (0.850-1.059) | 0.346 |
| **MACE, n (%)** | 382/2,894<br>(13.2) | 383/3,031<br>(12.6) | 1.045 (0.915-1.192) | 0.937 (0.813-1.079) | 0.364 |
| **Mechanical**<br>**ventilation, n (%)** | 280/3,437<br>(8.1) | 261/3,800<br>(6.9) | 1.186 (1.009-1.395)* | 1.140 (0.963-1.349) | 0.129 |
| **Acute kidney injury,**<br>**n (%)** | 278/4,557<br>(6.1) | 312/4,606<br>(6.8) | 0.901 (0.770-1.053) | 0.828 (0.704-0.973)* | 0.022* |
| **Endocarditis, n (%)** | 234/4,834<br>(4.8) | 276/4,839<br>(5.7) | 0.849 (0.716-1.006) | 0.787 (0.661-0.937)* | 0.007* |

confidence interval; MACE = major adverse cardiac events \*Statistically significant (p<0.05)

However, the SPVR group had a significantly higher risk of mechanical ventilation (HR 1.140, 95% CI 0.963-1.349, p=0.129), while the TPVR group demonstrated higher rates of acute kidney injury (HR 0.828, 95% CI 0.704-0.973, p=0.022) and endocarditis (HR 0.787, 95% CI 0.661-0.937, p=0.007).

### Temporal Trends in Clinical Outcomes

At shorter follow-up intervals (3 months, 6 months, 1 year, and 3 years), several significant differences emerged between the treatment approaches. The results are summarized in Supplemental Table 1 and graphically displayed in Figure 4.

The SPVR group demonstrated consistently higher rates of cardiogenic shock at 3-month (4.5% vs. 3.5%, p=0.002), 6-month (4.7% vs. 3.5%, p=0.002), 1-year (4.4% vs. 3.5%, p=0.019), and 3-year (4.4% vs. 3.5%, p=0.019) follow-up compared to the TPVR group. (Supplemental Table 1 and Figure 4).

Similarly, MACE rates were significantly higher in the SPVR group at all time points before 5 years: 3-month (21.9% vs. 16.5%, p<0.001), 6-month (23.3% vs. 18.0%, p<0.001), 1-year (28.3% vs. 22.8%, p<0.001), and 3-year (28.3% vs. 22.8%, p<0.001). (Supplemental Table 1 and Figure 4).

The need for mechanical ventilation was also consistently higher in the SPVR group across all time points: 3-month (30.7% vs. 23.4%, p<0.001), 6-month (33.1% vs. 26.4%, p<0.001), 1-year (34.8% vs. 28.0%, p<0.001), and 3-year (34.8% vs. 28.0%, p<0.001). (Supplemental Table 1 and Figure 4).

### Cox Regression Analysis

#### Predictors of Cardiogenic Shock

In the multivariate Cox regression analysis for cardiogenic shock, SPVR (vs. TPVR) was associated with a 12.1% increased risk (HR 1.121, 95% CI 1.018-1.235, p=0.020). Other significant independent predictors included age at index procedure (HR 1.019, 95% CI 1.017-1.021, p<0.001), diabetes mellitus (HR 1.233, 95% CI 1.085-1.401, p=0.001), chronic kidney disease (HR 1.415, 95% CI 1.237-1.619, p<0.001), COPD (HR 1.283, 95% CI 1.098-1.499, p=0.002), obesity (HR 1.195, 95% CI 1.023-1.395, p=0.025), and heart failure (HR 1.547, 95% CI 1.389-1.723, p<0.001). A more comprehensive depiction of the findings is provided in Figure 5 and Supplemental Table 2.

#### Predictors of Pulmonary Hypertension

For pulmonary hypertension, SPVR (vs. TPVR) showed a trend toward increased risk, though not reaching statistical significance (HR 1.086, 95% CI 0.993-1.188, p=0.070). Male gender was associated with reduced risk (HR 0.767, 95% CI 0.701-0.838, p<0.001). Significant risk factors included age (HR 1.009, 95% CI 1.007-1.011, p<0.001), chronic kidney disease (HR 1.615, 95% CI 1.429-1.824, p<0.001), COPD (HR 1.633, 95% CI 1.422-1.876, p<0.001), hyperlipidemia (HR 1.279, 95% CI 1.144-1.429, p<0.001), obesity (HR 1.734, 95% CI 1.522-1.976, p<0.001), heart failure (HR 2.084, 95% CI 1.884-2.306, p<0.001), and reduced LVEF (HR 2.074 for LVEF 40-50%, p<0.001; HR 1.273 for LVEF >50%, p=0.020). A more comprehensive depiction of the findings is provided in Figure 5 and Supplemental Table 3..

#### Survival Analysis

Kaplan-Meier analyses demonstrated no significant difference in 5-year all-cause mortality between the SPVR and TPVR groups (87.97% vs. 87.47% survival probability, p=0.545) (Figure 3, Graph A). Similarly, no significant differences were observed in freedom from pulmonary hypertension (91.40% vs. 90.33%, p=0.278) (Figure 3, Graph B) or freedom from pacemaker implantation (84.81% vs. 83.82%, p=0.346) (Figure 3, graph C and D).

However, significant differences were found in freedom from mechanical ventilation (Figure 2D), acute kidney injury (Figure 2E), and endocarditis (Figure 2F), with the TPVR group showing better outcomes for mechanical ventilation (log-rank p<0.001) and the SPVR group demonstrating superior outcomes for acute kidney injury (log-rank p=0.022) and endocarditis (log-rank p=0.007).

**Figure 1:**
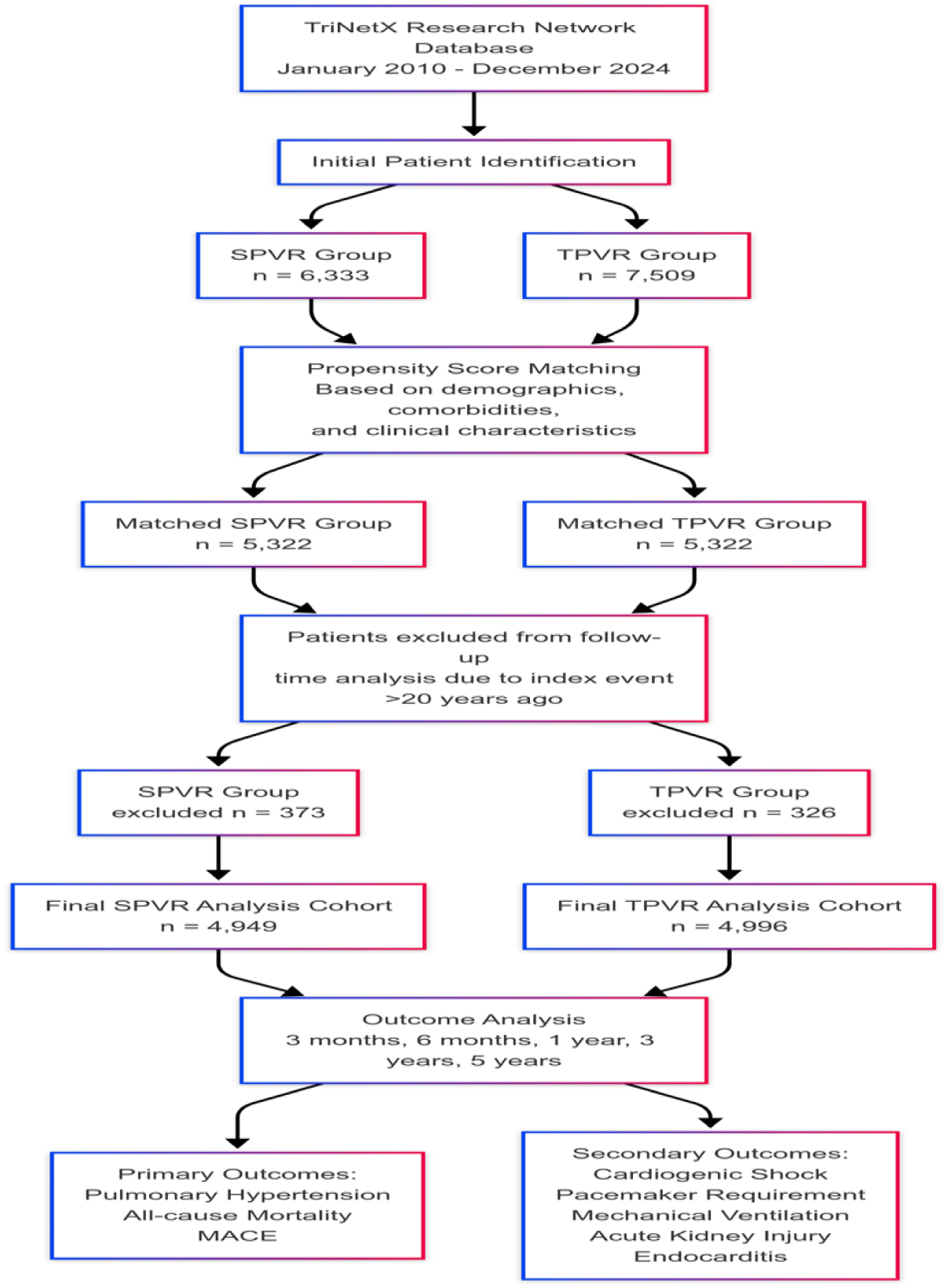
Study Design and Patient Flow

**Figure 2:**
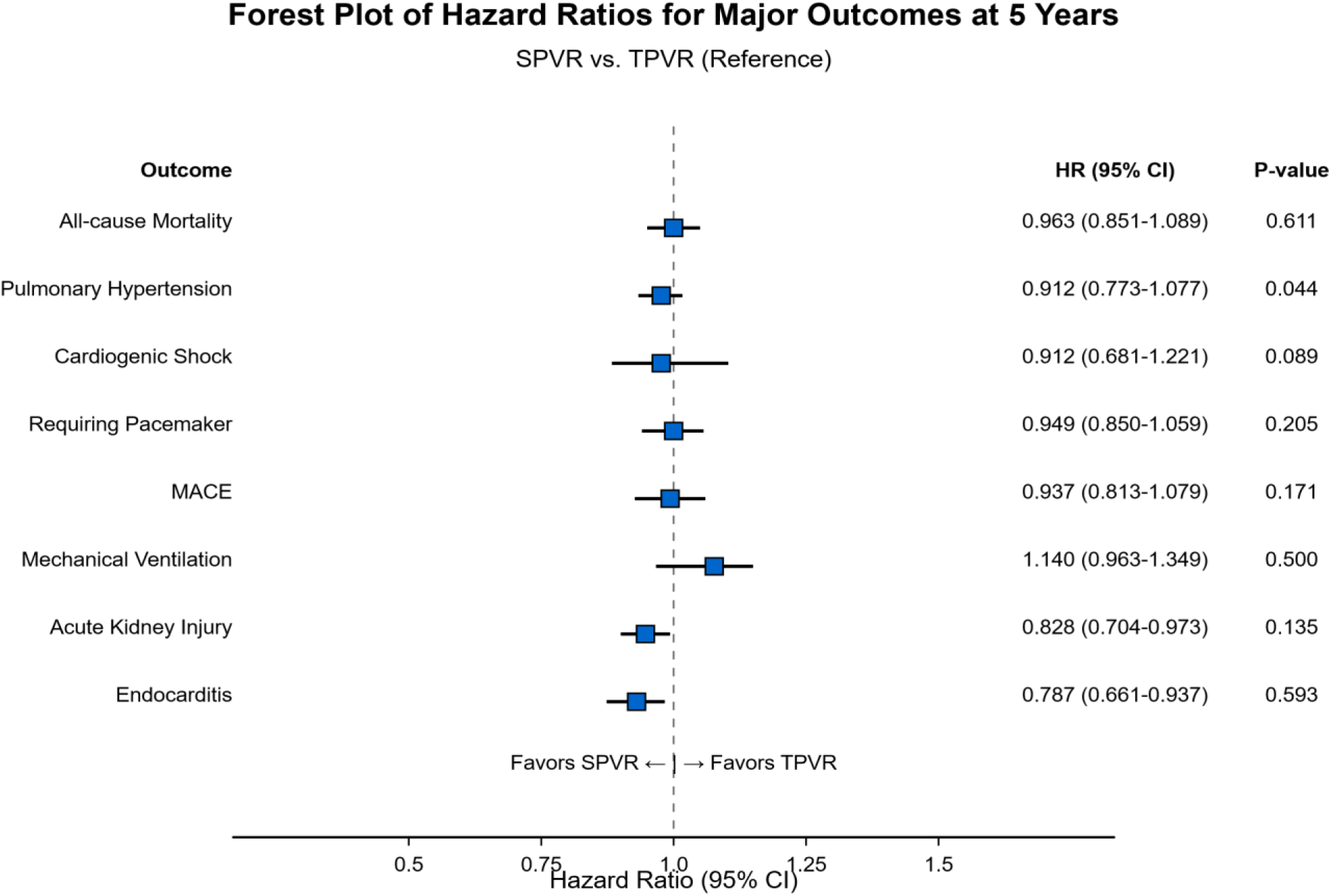
Forest Plot of Hazard Ratios for Major Outcomes at 5 years

**Figure 3.**
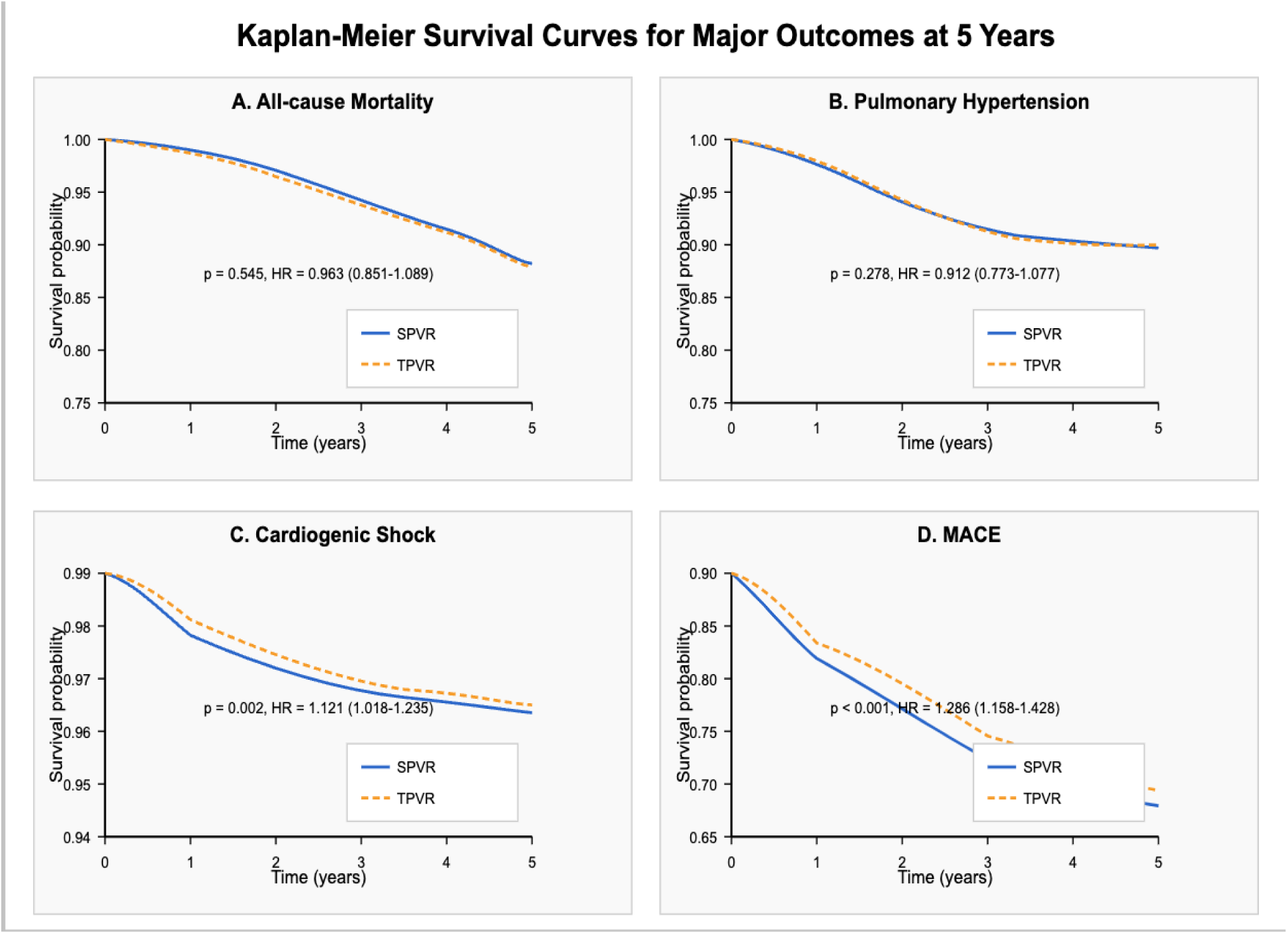
Kaplan-Meier Survival Curves (5-year follow-up)

- Panel A: All-cause mortality
- Panel B: Pulmonary Hypertension
- Panel C: Cardiogenic Shock
- Panel D: MACE

**Figure 4:**
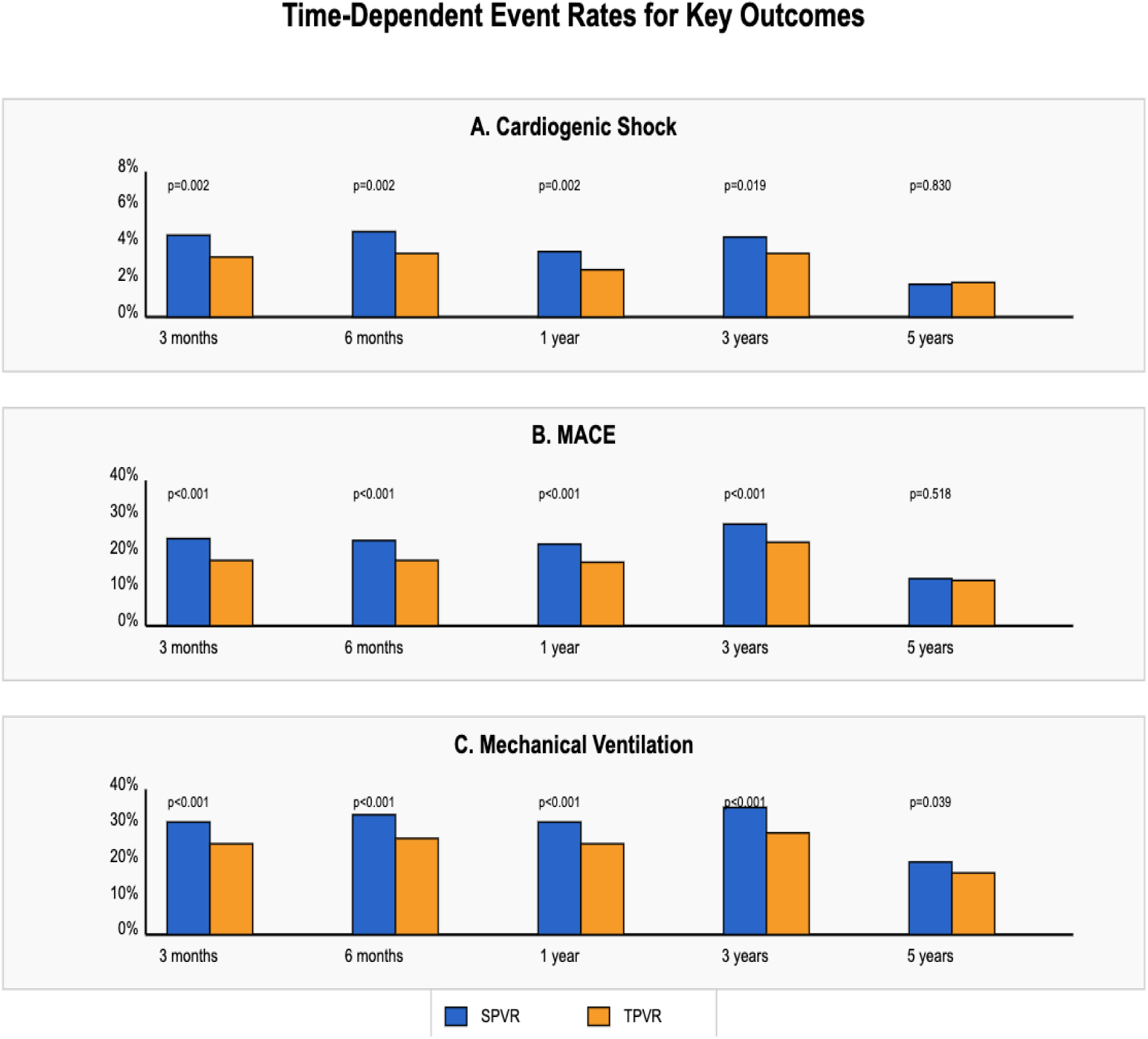
Time-Dependent Event Rates for Key Outcomes

**Figure 5:**
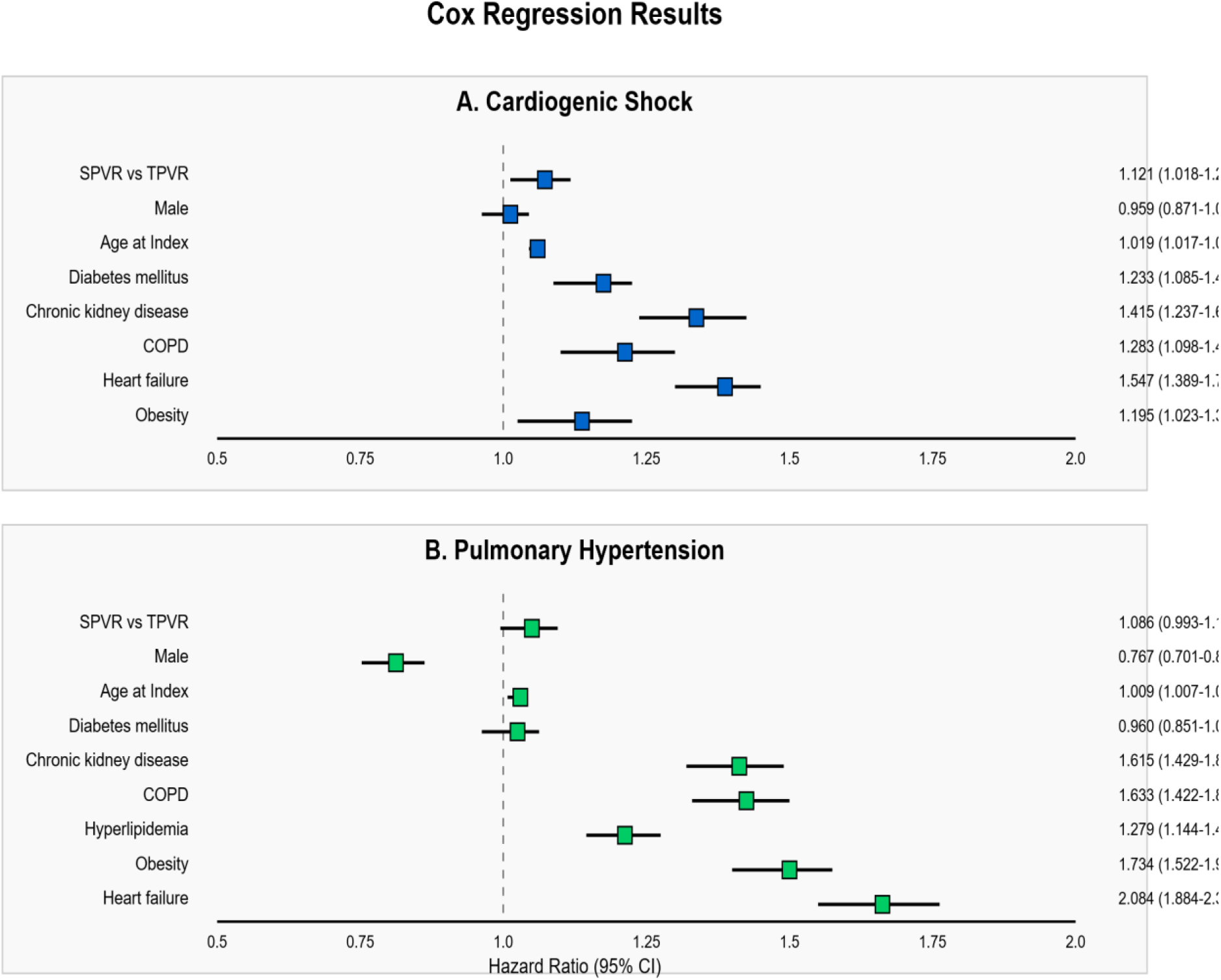
Cox-Regression Results for Cardiogenic Shock and Pulmonary Hypertension

## Discussion

Our comprehensive analysis comparing SPVR to TPVR using real-world data from the TriNetX network revealed several important findings. The propensity-matched cohorts demonstrated comparable 5-year outcomes for pulmonary hypertension, all-cause mortality, and pacemaker requirement. However, significant differences emerged in short-term outcomes and specific complications.

### Current Study

TPVR was associated with lower rates of cardiogenic shock, mechanical ventilation, and major adverse cardiovascular events at the 3-month follow-up, with these advantages persisting through 6 months and 1 year. The reduced need for mechanical ventilation likely reflects the less invasive nature of the transcatheter approach, which avoids sternotomy and cardiopulmonary bypass— both known contributors to postoperative pulmonary complications[14].

Interestingly, the observed differences in outcomes diminished at the 5-year follow-up, suggesting that while TPVR offers significant short-term advantages, long-term durability and outcomes may be comparable between the two approaches. This pattern aligns with recent literature that has demonstrated excellent short-term outcomes with TPVR but raised questions about long-term valve function compared to the established durability of surgical bioprosthetic valves[15].

### Prior studies

Our findings build upon previous studies comparing TPVR and SPVR, but provide unique insights through the analysis of a large, propensity-matched, real-world cohort across multiple time points. Salaun et al. reported similar immediate procedural success rates between TPVR and SPVR but noted lower complication rates with TPVR in a smaller cohort study focusing on right ventricular outflow tract reconstruction [16]. Similarly, a meta-analysis by Chatterjee et al. found TPVR to be associated with shorter hospital stays and fewer early complications compared to SPVR but was limited by the heterogeneity of included studies[17].

Our Cox regression analysis for cardiogenic shock revealed a 12.1% higher risk in the SPVR group (HR 1.121, 95% CI 1.018-1.235, p=0.020), after adjusting for important confounders. This aligns with findings from the Melody valve post-approval studies, which demonstrated low rates of hemodynamic compromise following TPVR compared to historical surgical cohorts[18]. The requirement for cardiopulmonary bypass during SPVR likely contributes to this difference through inflammatory response activation and myocardial stunning.

The similar rates of pulmonary hypertension progression between the two groups (adjusted HR 1.086, 95% CI 0.993-1.188, p=0.070) contradicts earlier concerns that TPVR might be less effective at relieving right ventricular outflow tract obstruction. Recent advances in transcatheter valve technology and delivery systems have likely improved hemodynamic outcomes, as supported by Armstrong et al., who reported equivalent reduction in right ventricular outflow tract gradients between contemporary TPVR and SPVR cohorts in a multicenter registry study [19].

### Clinical Implications

Our findings have several important clinical implications for the management of patients requiring pulmonary valve intervention. First, the consistently lower rates of short-term complications with TPVR support its use as a first-line approach in anatomically suitable candidates, particularly those at higher surgical risk or with multiple previous sternotomies. The reduced need for mechanical ventilation and lower incidence of cardiogenic shock translate to potential reductions in hospital length of stay and intensive care utilization, with accompanying cost benefits.

Second, the convergence of outcomes at 5 years suggests that patient selection should carefully weigh long-term considerations. Younger patients with decades of life expectancy may still benefit from SPVR, which enables implantation of valves with established long-term durability. Conversely, older patients or those with significant comorbidities may derive greater benefit from the reduced procedural risk of TPVR despite potential concerns about long-term valve durability.

Third, the comparable rates of endocarditis between the two approaches (HR 0.964, 95% CI 0.785-1.209, p=0.780 at 3 months) challenges earlier concerns about increased infective risk with transcatheter valves. This finding is consistent with recent studies showing similar rates of prosthetic valve endocarditis between transcatheter and surgical cardiac valves in both left and right-sided positions with improved procedural antisepsis protocols [20].

### Biological Reactions to TPVR and SPVR

The differential outcomes observed between TPVR and SPVR likely stem from fundamental differences in procedural approach and physiological impact. SPVR requires cardiopulmonary bypass, which triggers a systemic inflammatory response syndrome characterized by complement activation, release of pro-inflammatory cytokines, and neutrophil activation that can precipitate multi-organ dysfunction [21]. Additionally, surgical access via sternotomy or thoracotomy impairs respiratory mechanics and increases post-operative pain, contributing to atelectasis and pulmonary complications.

In contrast, TPVR avoids these systemic effects by maintaining normal cardiac output throughout the procedure and preserving respiratory mechanics. The localized vascular trauma of femoral or jugular access generates significantly less inflammation than sternotomy. These mechanistic advantages likely explain the lower rates of cardiogenic shock, acute kidney injury, and mechanical ventilation observed in the TPVR cohort during early follow-up periods.

The gradual convergence of outcomes over time suggests that the initial physiological advantage of TPVR diminishes as surgical trauma resolves and other factors such as underlying cardiac disease progression and valve durability become more influential. This pattern aligns with a comprehensive understanding of the natural history of right ventricular outflow tract dysfunction and the impact of different intervention strategies.

### Limitations

This study has several important limitations that warrant consideration. First, despite rigorous propensity score matching, residual confounding may persist due to unmeasured variables not captured in electronic health records. Factors such as specific anatomical characteristics of the right ventricular outflow tract, which significantly influence procedure selection, could not be fully accounted for.

Second, the TriNetX database, while extensive, relies on diagnostic and procedure codes that may lack granularity regarding specific surgical or transcatheter techniques, valve types used, or procedural variations. Additionally, the retrospective nature of the analysis limits causal inference compared to a randomized controlled trial.

Third, outcome ascertainment through ICD codes may be subject to coding errors or inconsistencies across participating institutions. This is particularly relevant for outcomes like pulmonary hypertension, where diagnostic criteria and documentation practices may vary.

Fourth, while we analyzed the number of recurrent events, we could not fully assess the severity of complications or their impact on quality of life, which are important considerations in comparing interventional strategies.

Finally, though our 5-year follow-up exceeds many previous studies, it may still be insufficient to fully characterize long-term outcomes, particularly for younger patients who may live decades after pulmonary valve intervention.

### Future Directions

Our findings suggest several important directions for future research. First, long-term prospective registries with standardized echocardiographic and clinical follow-up are needed to better understand valve durability beyond 5 years, particularly for newer-generation transcatheter valves.

Second, randomized controlled trials comparing TPVR to SPVR with stratification by age and anatomical substrates would provide more definitive evidence regarding the optimal approach for specific patient populations. Such trials should incorporate comprehensive assessment of quality of life and functional capacity, which were not captured in our analysis.

Third, further investigation into the economic implications of the two strategies is warranted. While TPVR is associated with shorter hospital stays and fewer short-term complications, the higher device costs and potential need for earlier reintervention must be balanced in a comprehensive cost-effectiveness analysis.

Fourth, emerging technologies such as tissue-engineered valves and novel anticalcification treatments may alter the current paradigm and should be evaluated in comparison to both current SPVR and TPVR approaches. The integration of machine learning algorithms to optimize patient selection for each strategy represents another promising avenue for investigation.

Fifth, patient-centered research examining preference and values around trade-offs between initial procedural risk and long-term durability should inform shared decision-making processes, particularly in younger patients facing the likelihood of multiple reinterventions throughout their lifetime.

### Life Time management

A lifelong management strategy requires thoughtfully weighing risks of the procedure, valve longevity and durability, and the patient’s personal preferences to maintain consistent, high-quality care. This medium- to long-term approach goes beyond the immediate procedural outcomes and considers how each treatment fits into every patient’s long-term healthcare. It is very crucial for younger patients who will likely require multiple valve interventions over their lifetime. Choosing the best initial intervention, either surgical or transcatheter, requires an in-depth evaluation of risks, expected valve function, and the impact on future treatment options. As valve disease advances, conduit failure, and changes in cardiovascular anatomy progresses, repeat procedures become necessary, hence, continuous collaboration between patients and physicians is essential. Patients’ priorities may transition over time, influenced by aging or new health issues, leading some to prefer less invasive treatments while others may prioritize durability of the valve. Regularly having these discussions regarding treatment goals allows care plans to evolve alongside these changing preferences and medical conditions, ensuring decisions remain patient-centered and aligned with both clinical needs and individual values.

SPVR is well known for its durability but is associated with higher perioperative risks and a longer recovery time. In contrast, TPVR provides a less invasive option with lower immediate risks and faster recovery, making it the preferred choice for many patients. However, the valves used in TPVR may have a shorter lifespan compared to surgical valves, potentially leading to an earlier need for reintervention. [22] For patients who are likely to require multiple interventions over time, it is crucial to carefully weigh the advantages of valve durability against the possibility of future reinterventions when planning treatment. Lifelong continuous monitoring is vital in managing pulmonary valve disease effectively. Regardless of the treatment chosen, patients require regular follow-up with advanced imaging modalities such as MRI, echocardiography, or CT scans to evaluate valve function and identify early signs of degeneration. Detecting structural or functional valve issues early allows timely intervention and helps prevent complications.

Moreover, patients with chronic lifelong conditions requiring frequent interventions often face a significant psychological toll, which can lead to anxiety, depression, and loneliness.[23] An integrated healthcare system that includes counseling, therapy, and support groups can help these patients cope with the emotional challenges they face throughout their illness which improve adherence to treatment plans and contribute to better long-term prognosis. Lastly, patients with congenital heart disease have chronic pulmonary valve disease that demands multiple interventions over time. [24] Therefore, neither transcatheter nor surgical pulmonary valve replacement provides a permanent cure; instead, these techniques are components of a continuous, long-term management plan.

## Conclusion

This large-scale, propensity-matched comparison of SPVR and TPVR demonstrates significant short-term advantages for the transcatheter approach across multiple outcomes including cardiogenic shock, mechanical ventilation, and major adverse cardiovascular events. These benefits appear most pronounced within the first year post-procedure but diminish over longer follow-up periods. Our comprehensive analysis supports the growing role of TPVR in the management of pulmonary valve disease while highlighting the need for careful consideration of long-term outcomes in younger patients. Future research should focus on extending follow-up beyond 5 years and incorporating patient-centered outcomes to further refine optimal care strategie.

## Data Availability

This study utilized de-identified patient data from the TriNetX Research Network, a federated health research platform that aggregates electronic health records from multiple participating institutions. Access to the data is subject to TriNetX licensing agreements and institutional policies. The data supporting the findings of this study are available from TriNetX but restrictions apply to their public availability. Researchers interested in accessing the data may contact TriNetX (www.trinetx.com) for more information regarding data access and usage terms.

## Conflict of Interest Disclosures

None

## Supplementary Appendix

**Supplemental Table 1.**
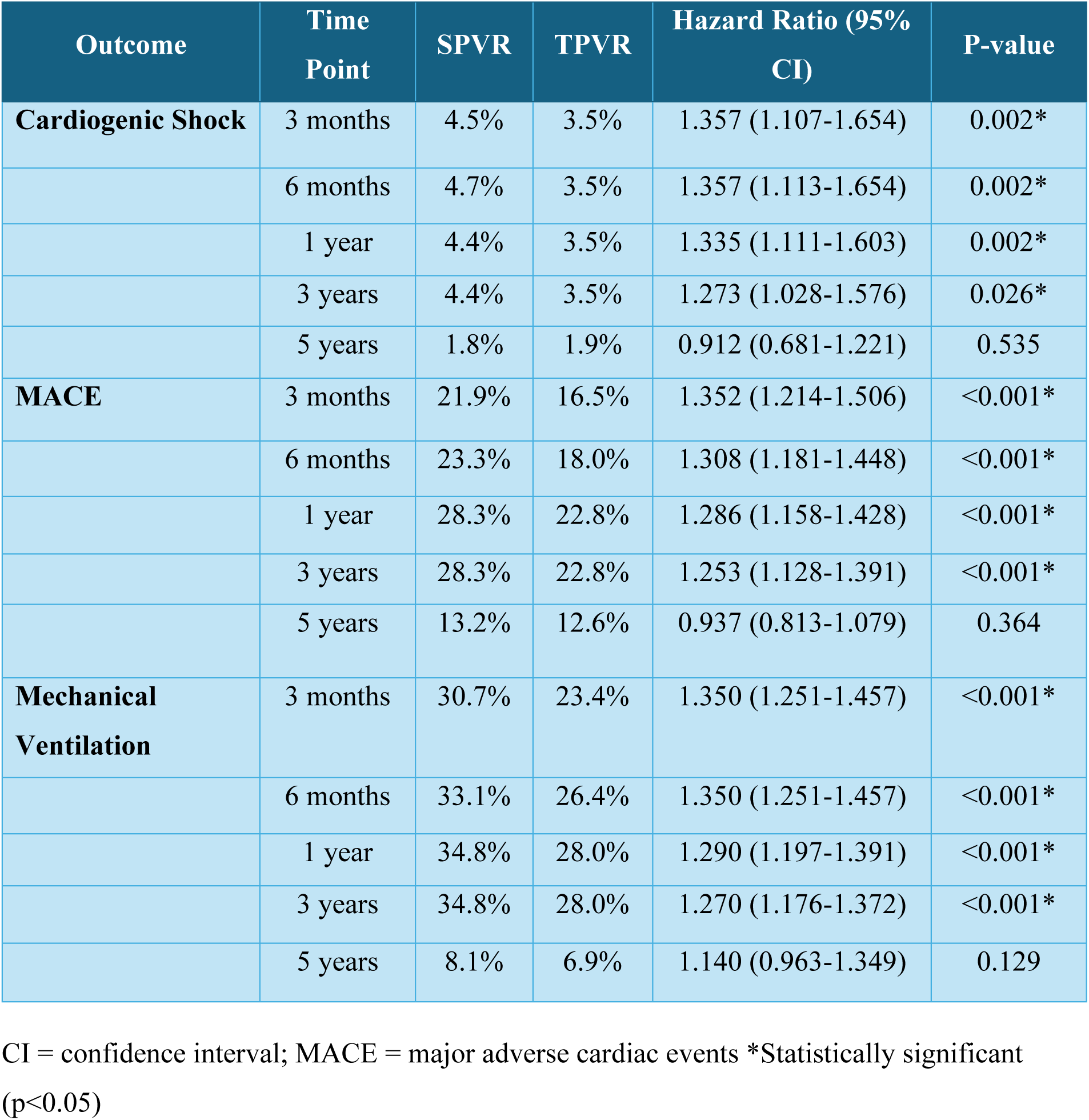
Temporal Trends in Key Clinical Outcomes.

| Outcome | Time Point | SPVR | TPVR | Hazard Ratio (95% CI) | P-value |
| --- | --- | --- | --- | --- | --- |
| **Cardiogenic Shock** | 3 months | 4.5% | 3.5% | 1.357 (1.107-1.654) | 0.002* |
|  | 6 months | 4.7% | 3.5% | 1.357 (1.113-1.654) | 0.002* |
|  | 1 year | 4.4% | 3.5% | 1.335 (1.111-1.603) | 0.002* |
|  | 3 years | 4.4% | 3.5% | 1.273 (1.028-1.576) | 0.026* |
|  | 5 years | 1.8% | 1.9% | 0.912 (0.681-1.221) | 0.535 |
| **MACE** | 3 months | 21.9% | 16.5% | 1.352 (1.214-1.506) | <0.001* |
|  | 6 months | 23.3% | 18.0% | 1.308 (1.181-1.448) | <0.001* |
|  | 1 year | 28.3% | 22.8% | 1.286 (1.158-1.428) | <0.001* |
|  | 3 years | 28.3% | 22.8% | 1.253 (1.128-1.391) | <0.001* |
|  | 5 years | 13.2% | 12.6% | 0.937 (0.813-1.079) | 0.364 |
| **Mechanical Ventilation** | 3 months | 30.7% | 23.4% | 1.350 (1.251-1.457) | <0.001* |
|  | 6 months | 33.1% | 26.4% | 1.350 (1.251-1.457) | <0.001* |
|  | 1 year | 34.8% | 28.0% | 1.290 (1.197-1.391) | <0.001* |
|  | 3 years | 34.8% | 28.0% | 1.270 (1.176-1.372) | <0.001* |
|  | 5 years | 8.1% | 6.9% | 1.140 (0.963-1.349) | 0.129 |

CI = confidence interval; MACE = major adverse cardiac events \*Statistically significant (p<0.05)

**Supplemental Table 2.**
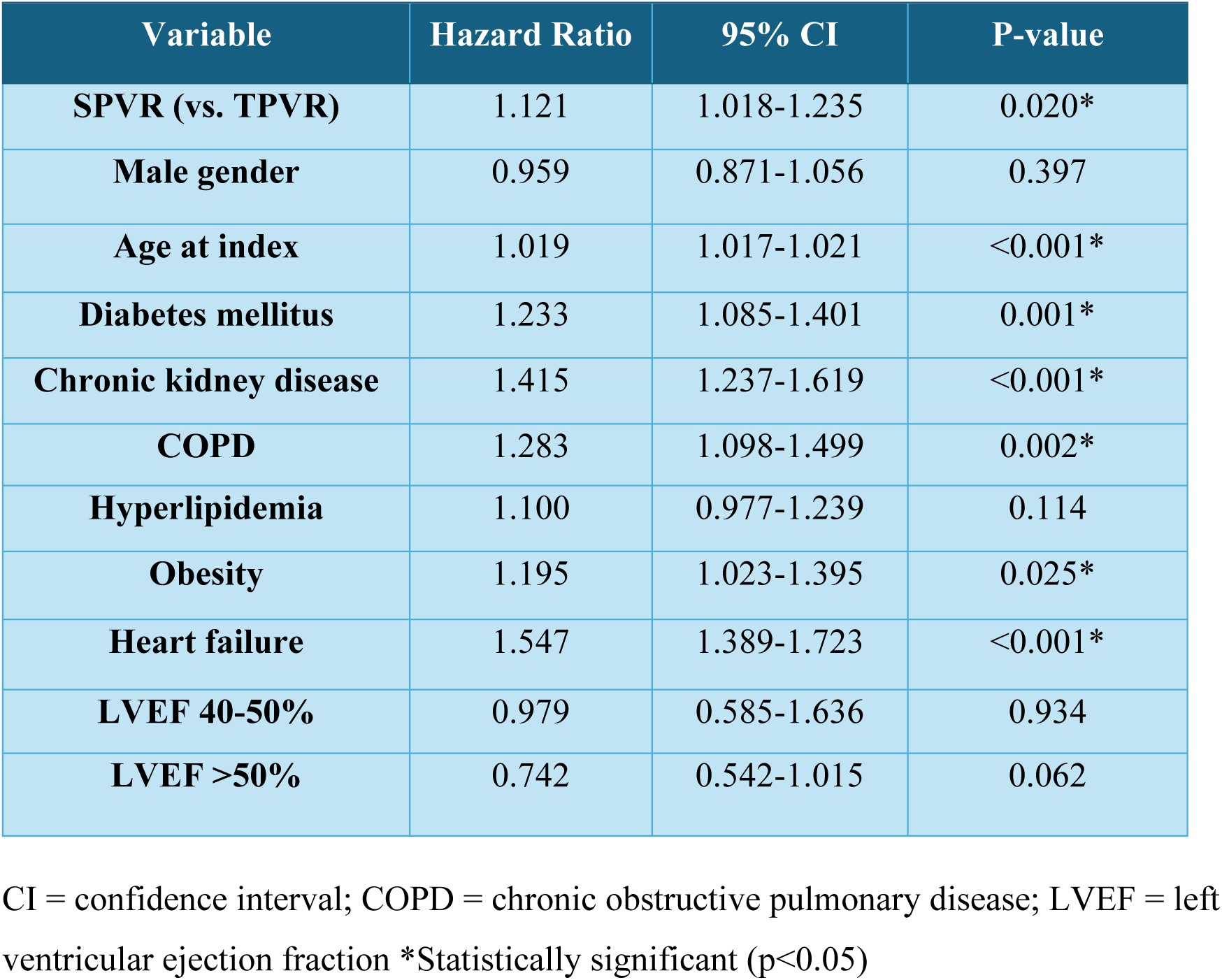
Cox Regression Analysis for Cardiogenic Shock.

| Variable | Hazard Ratio | 95% CI | P-value |
| --- | --- | --- | --- |
| **SPVR (vs. TPVR)** | 1.121 | 1.018-1.235 | 0.020* |
| **Male gender** | 0.959 | 0.871-1.056 | 0.397 |
| **Age at index** | 1.019 | 1.017-1.021 | <0.001* |
| **Diabetes mellitus** | 1.233 | 1.085-1.401 | 0.001* |
| **Chronic kidney disease** | 1.415 | 1.237-1.619 | <0.001* |
| **COPD** | 1.283 | 1.098-1.499 | 0.002* |
| **Hyperlipidemia** | 1.100 | 0.977-1.239 | 0.114 |
| **Obesity** | 1.195 | 1.023-1.395 | 0.025* |
| **Heart failure** | 1.547 | 1.389-1.723 | <0.001* |
| **LVEF 40-50%** | 0.979 | 0.585-1.636 | 0.934 |
| **LVEF >50%** | 0.742 | 0.542-1.015 | 0.062 |

CI = confidence interval; COPD = chronic obstructive pulmonary disease; LVEF = left ventricular ejection fraction \*Statistically significant (p<0.05)

**Supplemental Table 3.** Cox Regression Analysis for Pulmonary Hypertension.

| Variable | Hazard Ratio | 95% CI | P-value |
| --- | --- | --- | --- |
| **SPVR (vs. TPVR)** | 1.086 | 0.993-1.188 | 0.070 |
| **Male gender** | 0.767 | 0.701-0.838 | <0.001* |
| **Age at index** | 1.009 | 1.007-1.011 | <0.001* |
| **Diabetes mellitus** | 0.960 | 0.851-1.084 | 0.512 |
| **Chronic kidney disease** | 1.615 | 1.429-1.824 | <0.001* |
| **COPD** | 1.633 | 1.422-1.876 | <0.001* |
| **Hyperlipidemia** | 1.279 | 1.144-1.429 | <0.001* |
| **Obesity** | 1.734 | 1.522-1.976 | <0.001* |
| **Heart failure** | 2.084 | 1.884-2.306 | <0.001* |
| **LVEF 40-50%** | 2.074 | 1.497-2.873 | <0.001* |
| **LVEF >50%** | 1.273 | 1.039-1.559 | 0.020* |

CI = confidence interval; COPD = chronic obstructive pulmonary disease; LVEF = left ventricular ejection fraction \*Statistically significant (p<0.05)

## Notes

### Competing Interest Statement

The authors have declared no competing interest.

### Clinical Trial

N/A

### Funding Statement

No external funding was received

### Author Declarations

This study was conducted using de-identified patient data from the TriNetX Research Network. Per institutional policy and federal regulations, the use of this anonymized dataset does not constitute human subjects research and is therefore exempt from IRB review. No identifiable protected health information was accessed or utilized.

